# Hardware-Free Testing for Antimicrobial Resistance Using Artificial Intelligence

**DOI:** 10.1101/2024.07.11.24309858

**Authors:** Purbali Chakraborty, Mert Tunca Doganay, Abdullah Tozluyurt, Andrea M. Hujer, Robert A. Bonomo, Mohamed S. Draz

**Affiliations:** Department of Medicine, Case Western Reserve University School of Medicine, Cleveland, OH, 44106, USA; Research Service, Louis Stokes Cleveland Department of Veterans Affairs Medical Center, Cleveland, OH, USA; Departments of Molecular Biology and Microbiology, Biochemistry, Pharmacology, and Proteomics and Bioinformatics, Case Western Reserve University School of Medicine, Cleveland, OH, USA; CWRU-Cleveland VAMC Center for Antimicrobial Resistance and Epidemiology (Case VA CARES) Cleveland, OH, USA; Department of Biomedical Engineering, Case Western Reserve University, Cleveland, OH, USA; Department of Biomedical Engineering, Cleveland Clinic, Cleveland, OH, 44106, USA

## Abstract

Antimicrobial resistance (AMR) is one of the most challenging public health problems, and implementation of effective and accessible testing solutions is an ever-increasing unmet need. Artificial intelligence (AI) offers a promising avenue for enhanced testing performance and accuracy. We introduce an AI system specifically designed for rapid AMR testing, eliminating the requirement for bulky hardware and extensive automation. Our system incorporates a novel approach for nanotechnology-empowered intelligent diagnostics (NEIDx), leveraging nanoparticles to enable novel AI-based advanced systems for detection. We employ catalytic nanoparticle-based NEIDx coupled with magnetic separation to facilitate the direct detection of AMR-associated enzymes from blood samples. This is achieved through the formation of easily visible and detectable large bubbles, a process streamlined by AI running on a cellphone. We evaluated the performance of our AI system using two clinically relevant AMR enzymes: Klebsiella pneumoniae carbapenemase-2 (KPC-2) and Sulfhydryl variable-1 (SHV-1) β-lactamases. The system demonstrated qualitative detection with a sensitivity of 82.61% (CI of 79.7 - 85.5%) and a specificity of 92.31% (CI of 90.3 - 94.3%) in blood samples, respectively. This innovative approach holds significant promise for advancing point-of-care diagnostics and addressing the urgent need for rapid and accessible AMR testing in diverse healthcare settings.

## INTRODUCTION

The global challenge of antimicrobial resistance (AMR) stands as one of the most pressing threats to public health, contributed to by the widespread and inappropriate use of antibiotics in both healthcare and livestock industries[1-3]. Beyond compromising the efficacy of potent antibiotics, the rapid evolution of resistant bacteria, either through intrinsic mechanisms or genetic alterations, coupled with the horizontal transfer of AMR genes among diverse strains, presents profound challenges in clinical settings. Therefore, the imperative for rapid and accessible AMR testing has become paramount to safeguard public health on a global scale[4, 5].

Artificial intelligence (AI) has emerged as a transformative force, revolutionizing various facets of healthcare[6]. Its application spans from clinical prognosis to direct-to-consumer testing, confirming its potential to address challenges in diverse medical fields. AI systems can be trained to discern pathological conditions with similar symptoms, enabling accurate clinical diagnosis[6-8]. This paradigm shift is exemplified by recent studies where AI has demonstrated superiority in biosensing, imaging, and medical diagnostics, showcasing its transformative impact across diverse healthcare domains[9-14]. The integration of AI and point-of-care (POC) testing marks a pivotal advancement. Numerous studies have successfully utilized AI for POC testing, enhancing the accuracy and efficiency of diagnostics[15-17]. However, despite these advancements, challenges persist in ensuring seamless and reliable testing without the involvement of healthcare professionals or the need of expensive instrumentation, thereby ensuring broad accessibility[18].

When combined with other innovative approaches, such as nanoparticle engineering, AI presents an opportunity for robust and hardware-free systems that is proper for POC testing in resource-constrained settings. Engineered nanoparticle systems, including gold, silver, platinum, and various other metallic and non-metallic nanoparticles, offer unique sensing properties that have found applications in AMR testing[19-24]. These nanomaterials, due to their distinctive physicochemical characteristics, play a significant role in enhancing the sensitivity and specificity of diagnostic assays, contributing to the efficacy and precision of biological detection methodologies[25-28]. Studies have shown the feasibility of AI and nanoparticles in enhancing AMR testing results. One key finding across these studies is the development of novel biosensors utilizing nanoparticles as sensing elements. These biosensors leverage the unique properties of nanoparticles, such as high surface area-to-volume ratio and tunable surface chemistry, to detect resistant bacterial strains with high sensitivity and selectivity. Machine learning and deep learning algorithms were integrated with silver nanoparticles[29-33], silicon nanowires[30], gold nanoparticles and nanowires[34-37], and gold-silver bimetallic nanostructures[38] to analyze the complex data generated by surface enhanced Raman spectroscopy (SERS) technique, enabling rapid and accurate identification of antimicrobial-resistant pathogens. Wang et al, developed a machine-learning-aided cocktail assay with lanthanide nanoparticles, rapidly detecting biofilms and showing promise for eradicating them with antibiotic-loaded probes [39]. Yu et al, introduced a rapid technique utilizing plasmonic nanosensors and machine learning to identify antibiotic resistance in ESKAPE pathogens with high accuracy in under 20 minutes [40]. These innovative approaches hold significant potential for clinical applications in biomedical settings.

In this landscape, we developed NEIDx as a novel system for rapid and highly accessible AMR testing. Our approach harnesses the power of AI to perform deep and accurate image analysis of bubble signal generated by a platinum-based catalytic nanoparticle system engineered to eliminate the need for extensive instrumentation. The developed NEIDx system allows for rapid (<60 minutes) detection of the targeted AMR-associated enzymes with 83% sensitivity and 92% specificity. This innovative, simple system not only ensures advanced and enhanced diagnostics but also operates without the need for specialized hardware, making it an ideal POC testing solution for accessible AMR testing in diverse settings, particularly resource-limited settings. Our system promises precision and efficiency in diagnostics, and also represents a transformative stride in addressing the global menace of antimicrobial resistance.

## RESULTS AND DISCUSSION

### NEIDx system development and characterization

The NEIDx system developed is a catalytic nanoparticle-empowered AI system that detects gas bubbles specifically formed and accumulated in the presence of an AMR-associated enzyme, including KPC-2 and SHV-1. NEIDx design relies on two major components for sample testing: (***i***) magnetic microbeads loaded with antibody to capture the target AMR enzyme, and (***ii***) Pt-nanoprobes prepared with catalytically active PtNPs, surface functionalized with target specific antibody to label the captured enzyme on the bead, inducing the formation of bubbles in the presence of a peroxide solution that can be detected using a simple optical system with a camera (e.g., a cellphone) (**Figure 1**).

**Figure 1.**
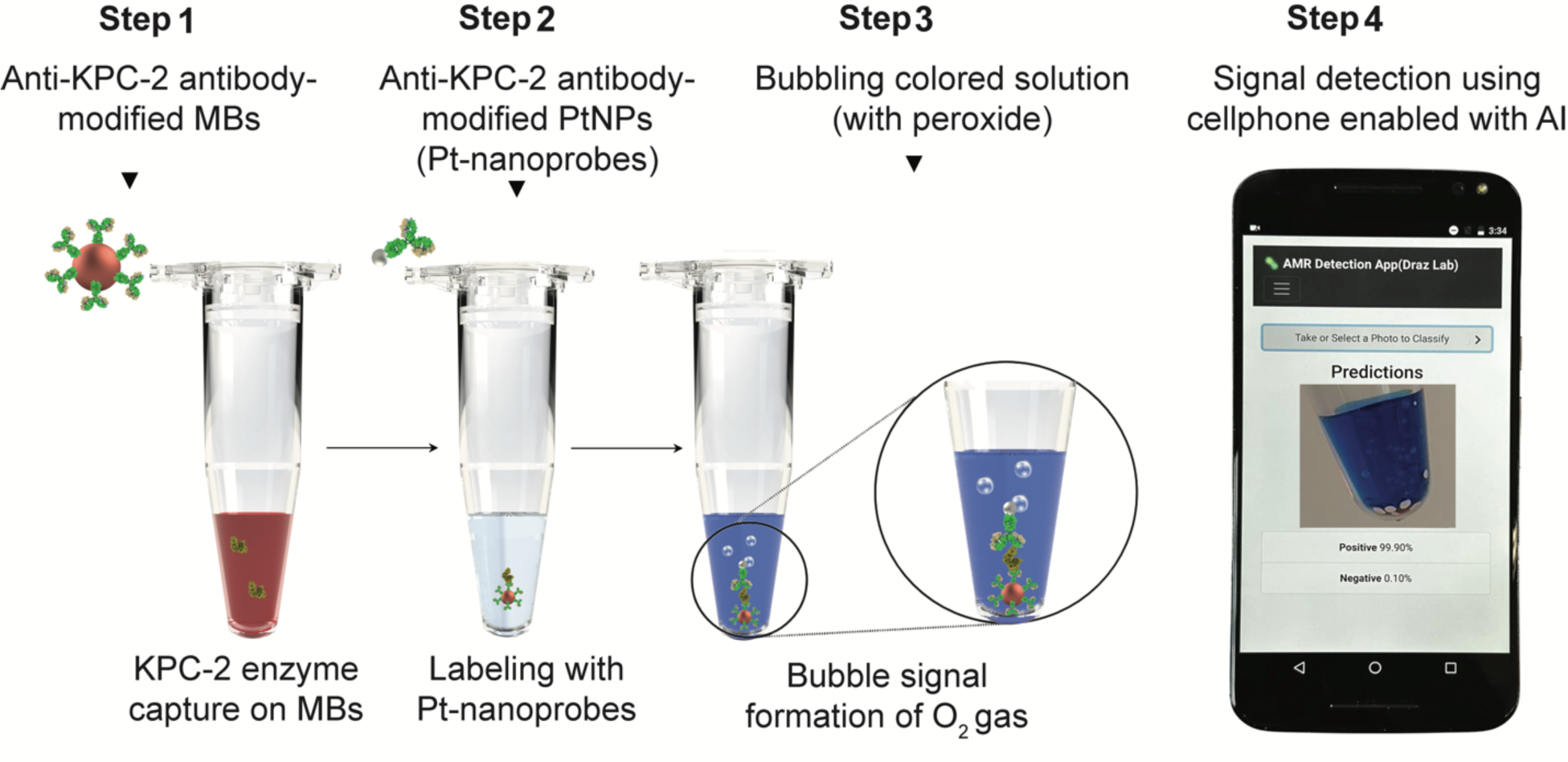
Schematic presentation of the NEIDx system. The developed system integrates catalytic nanoparticle-based NEIDx with magnetic separation to facilitate AMR testing using the following protocol: **Step 1** involves the capture of AMR enzymes on magnetic beads (MBs). Anti-KPC-2 antibody-modified MBs are introduced to the sample and allowed to incubate for 15 minutes at room temperature, facilitating the capture and isolation of KPC-2 enzyme on the surface of the beads; **Step 2** entails the labeling of AMR enzymes with catalytically-active nanoparticle probes. Washed MBs carrying the captured KPC-2 enzyme are incubated with Pt nanoprobes, which are prepared with platinum nanoparticles (PtNPs) surface-functionalized with anti-KPC-2 antibody. This process results in the formation of a three-component PtNP-KPC-2-MB complex; **Step 3** focuses on bubble signal formation. A peroxide-containing colored solution is added to the formed complex with PtNPs, initiating bubble signal formation; **Step 4** involves bubble imaging and signal detection using AI running on a cellphone, providing a streamlined and efficient approach to AMR detection.

Magnetic beads (MBs) coated with anti-KPC-2 and anti-SHV-1 antibody were employed to facilitate specific capture and isolation of the targeted AMR-associated β-lactamases of KPC-2 and SHV-1, respectively. The surface modification of the MBs was performed using a coupling reaction that allows for the directional conjugation of antibodies to the surface of the MB using one-step crosslinking chemistry. The reaction relies on a heterobifunctional crosslinker of 3-(2-pyridyldithio)propionyl hydrazide (PDPH) that contains sulfhydryl-reactive pyridyldithiol and carbonyl-reactive hydrazide moieties. Thiol-activated MBs with a size of 5 μm were first incubated with PDPH to prepare hydrazide-activated MBs with free hydrazide groups on their surface. Oxidized KPC-2 and SHV-1 antibodies—with Fc regions carrying free aldehyde groups, were then coupled to the surface of beads, forming highly reactive antibody-MB conjugates (**Figure 2a**). The prepared conjugates were tested using UV−vis spectroscopy. Results demonstrated the presence of a robust peak at 278 nm that can be attributed to KPC-2 antibody on the surface of the MBs, which was additionally confirmed by Fourier transform-infrared spectroscopy (FT-IR) analysis that showed a cluster of characteristic peaks at 1042.3, 1406.8, 1471.4, 1533.1, 1633.4, 2165.7, 2324.7, 2361.4, 2956.3, 3248.5, and 3327.6 cm^−1^. The peaks at 1533.1, 1633.4, and 3327.6 cm^−1^ can be attributed to the amide-I and II characteristic of the conjugated anti-KPC-2 antibody as protein structure, while the peaks at 3248.5 and 2956.8 cm^−1^ can be attributed to C=O of the carboxyl groups in the PDPH crosslinker utilized in the conjugation protocol (**Figure 2c and Table S1**)[41-43]. Laser scanning confocal microscopy (LSCM) analysis of MBs-coated with anti-KPC-2-Ab counterstained with Protein G-FITC also showed a successful coupling of antibody to beads as observed by the strong green fluorescence observed due to FITC, with 81.96% increase in the fluorescence signal intensity when compared with control samples of unmodified MBs (**Figure 2d**). The surface modification of MBs with antibodies and their efficiency in KPC-2 capture were verified using sodium dodecyl-sulfate polyacrylamide gel electrophoresis (SDS-PAGE) technique, where the conjugated anti-KPC-2 antibodies showed IgG characteristic bands at 149.34 KDa and 53.51 KDa and the captured KPC-2 on the surface of MB indicated the presence of an intense band at 24.86 KDa that is characteristic of the KPC-2 β-lactamase, confirming highly stable binding and effective isolation of the targeted enzyme molecules on the surface of MBs conjugates (**Figure 2e**)[27, 44].

**Figure 2.**
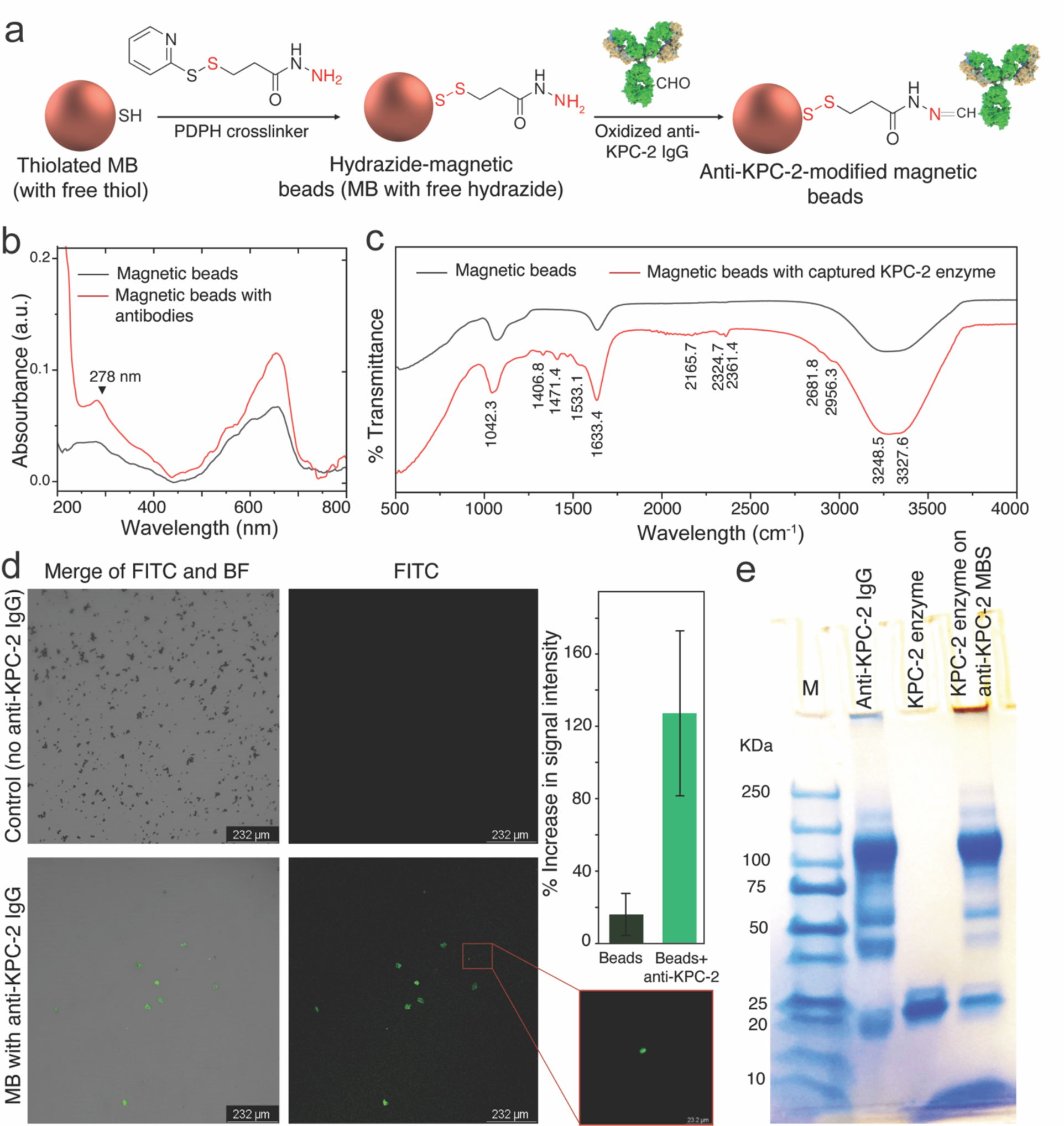
NEIDx magnetic beads preparation and characterization. (**a**) Schematic of magnetic bead (MB) surface modification with anti-KPC-2 antibody. MBs (5 µm) that carry thiol groups on its surface are initially activated with an excess of 3-(2-pyridyldithio)propionyl hydrazide (PDPH) crosslinker to prepare hydrazide-modified beads. Oxidized anti-KPC-2 antibodies with free aldehyde groups are coupled with the hydrazide groups presented on the surface of the beads following well-known Aldehyde-hydrazide coupling chemistry. (**b**) UV-vis absorbance spectra of the unmodified and anti-KPC-2 modified magnetic beads. (**c**) FT-IR analysis of unmodified and anti-KPC-2 modified magnetic beads. (**d**) Fluorescence spectroscopy analysis of unmodified and anti-KPC-2 modified magnetic beads stained with FITC conjugated protein G. The inset bar chart shows the increase in fluorescent signal intensity after bead surface modification with anti-KPC-2. (**e**) SDS-PAGE analysis confirming the presence of anti-KPC-2 antibody on the surface of beads and effective capture and isolation of the targeted KPC-2 enzyme.

Pt-nanoprobes were synthesized by functionalizing citrate-stabilized PtNPs with anti-KPC-2 or anti-SHV-1 antibodies (mAb) in accordance with our established surface chemistry protocol. This protocol relies on the utilization of PDPH to bind oxidized antibodies to the nanoparticle surface through their carbohydrate residues[45]. **Figure 3a** shows a digital image of citrate-stabilized PtNPs used in our protocol with its characteristic brown color. Transmission electron microscopy (TEM) and the corresponding size distribution histogram reveal that the PtNPs exhibit a spherical morphology, with an average diameter of 6.7 ± 2.318 nm (**Figure 3b, c**). Agarose gel electrophoresis, UV-vis and FT-IR spectroscopy techniques were employed for PtNPs surface chemistry and antibody immobilization characterization. Conjugation of antibodies to the PtNP surface led to retardation in the motion of the resulting Pt-nanoprobes (PtNPs-anti-KPC-2 Abs) compared to non-modified PtNPs during agarose gel electrophoresis. This retardation is attributed to differences in size and charge density values between PtNPs (no antibodies) and the formed nanoprobes (**Figure 3d**). In addition, the conjugation of mAb to PtNPs resulted in distinct peaks in the FT-IR analysis, indicative of antibody presence. In **Figure 3c**, FT-IR spectra of Pt-nanoprobes display bands at 3285.1, 2880.2, 1593.8, 1462.7, and 1403.9 cm^−1^, corresponding to N−H stretching, C=O stretching, N−H bending, C−H stretching and C−C stretching, respectively, confirming the stable coupling of anti-KPC-2 antibody to the surface of PtNPs[46, 47]. UV−Vis analysis of citrate-stabilized PtNPs and Pt-nanoprobes (PtNPs modified with anti-KPC-2 mAb) confirmed the stability of the synthesized nanoprobes. A strong absorption peak at 223 nm was observed, attributed to the presence of the antibody as a protein structure (**Figure 2e**). The antibody molecules per nanoparticle ratio was estimated to be 0.972 ± 0.809 antibody molecule/PtNP based on their absorption values at 223 nm. Consequently, approximately 3.46% of the Pt-nanoprobe particle surface was covered with anti-KPC-2 mAb, leaving 96.54% available for interaction with the peroxide containing-bubble solution. This antibody surface coverage ratio on Pt-nanoprobes facilitates efficient labeling of captured AMR enzymes (by increasing the number of PtNPs per captured KPC-2 enzyme on beads), minimizing the likelihood of large aggregate formation that could hinder bubble generation and lead to a false negative signal.

**Figure 3.**
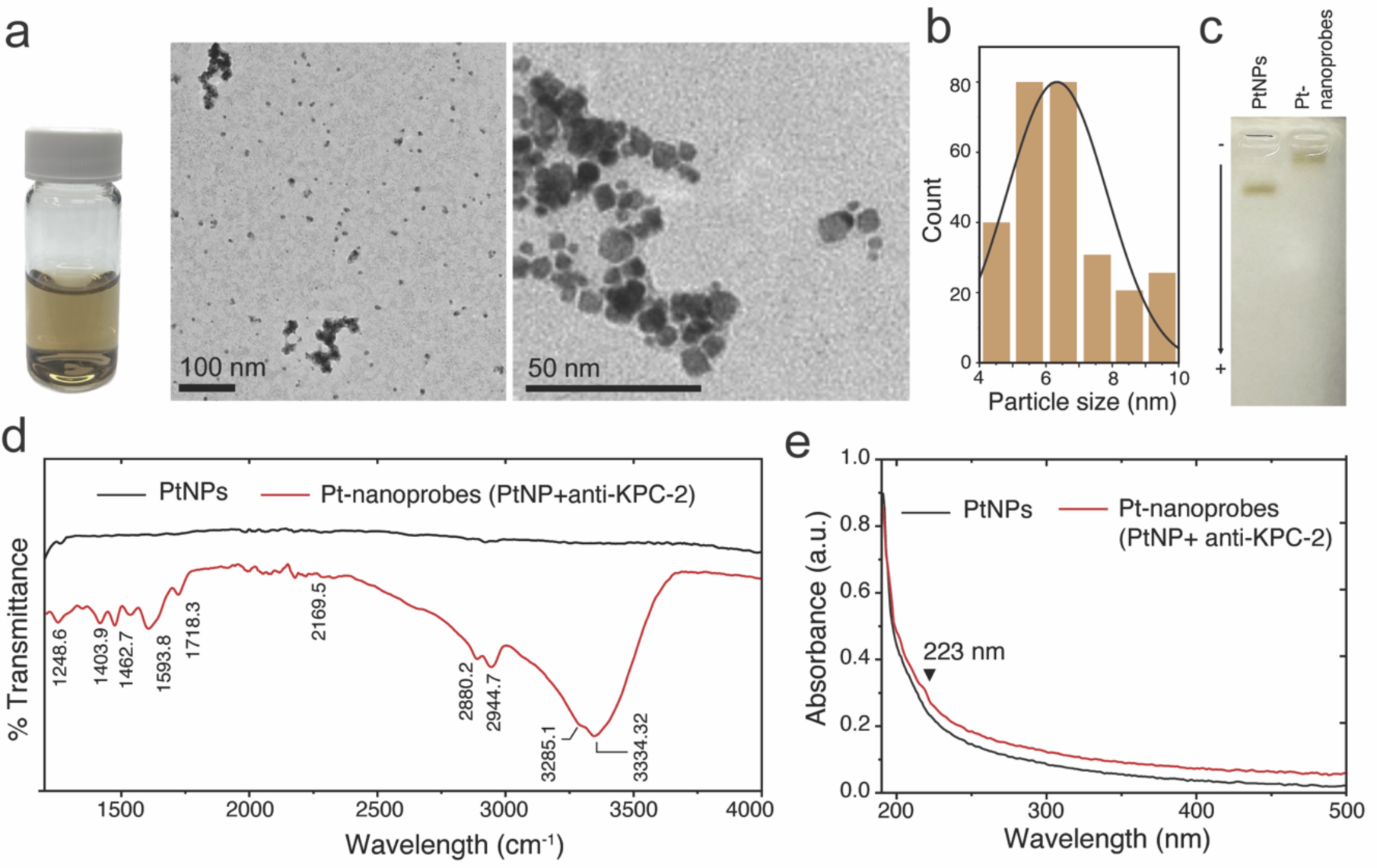
NEIDx Pt-nanoprobe preparation and characterization. (a) Digital image of PtNP (brown in color) solution utilized for Pt-nanoprobe preparation and the corresponding transmission electron microscopy micrographs of the as-prepared nanoparticle solution. (b) Particle size distribution histogram of the as-prepared PtNPs. (c) Agarose gel electrophoresis of PtNPs and anti-KPC-2 modified PtNPs (Pt-nanoprobes). (d) FT-IR analysis of unmodified and anti-KPC-2 modified PtNPs. (e) UV-vis spectroscopy analysis of the unmodified and anti-KPC-2 modified PtNPs.

### NEIDx system testing and validation

We started the testing process of the NEIDx system by first evaluating the bubble formation in the presence and absence of the targeted AMR enzymes, using KPC-2 as a model target. Then we tested the change in number of bubbles in the presence of serial concentrations of KPC-2 enzyme to confirm the efficiency of the proposed testing scheme and its sensitivity. Aliquots of KPC-2-spiked phosphate buffer (PB) with KPC-2 concentrations in a range from 0 – 10 ng/ml were added to anti-KPC-2 modified-MBs followed by the addition of Pt-nanoprobes (i.e., PtNPs modified with anti-KPC-2 antibodies). The formed PtNP-KPC-2-MB complexes were then added to a peroxide solution, containing 5% H₂O₂, and the generated bubbles were analyzed manually using ImageJ software (**Figure S1**). We noticed the formation of visible bubbles within 30 seconds, and the number of bubbles continued to increase with the incubation time (up to 2 – 5 minutes). The formed bubble signal showed a direct correlation with an increase in the concentration of the tested KPC-2 enzyme, demonstrating sensitivity down to 0.1 ng/ml (**Figure 4a and Figures S 2-4**). The entire testing protocol, from sample addition to the formation of bubbles, was completed within a mere 35 minute time frame. In addition to testing various concentrations of SHV-1 to confirm the efficiency of the developed testing protocol, we examined the flexibility for expanding the NEIDx testing scheme to include other targets, which is a key requirement for effective AMR testing and detection (**Figure 4b and Figures S 5-7**). The images presented in **Figure 4c** illustrate a direct increase in the bubbles generated to the increased concentration of the tested AMR KPC-2 (depicted in the top panel) and SHV-1 β-lactamases (depicted in the bottom panel). Using MBs modified with a mixture of anti-KPC-2 and anti-SHV-1 antibodies we were able to test samples that contain both types of β-lactamases. Samples spiked with both KPC-2 and SHV-1 at concentration of 5 ng/ml were incubated with MBs in the presence of a mixture of anti-KPC-2 and anti-SHV-1 Ab-modified Pt-nanoprobes (at 1:1 ratio). The generated bubble signal exhibited statistical significance (n = 3; *p* ≤ 0.05, employing paired Student’s t-test) in comparison to the control samples (no enzymes added), and the generated bubble signal surpassed that of each enzyme when tested individually (**Figure 4d and Figure S8**). The images presented in **Figure 4d and Figures S9-11** are representative of the tested samples containing KPC-2, SHV-1, or a combination of both enzymes, all at a concentration of 5 ng/ml. This substantiates the potential of the developed system to effectively test multiple target enzymes, even when dealing with closely related ones, where cross-reactivity might be common.

**Figure 4.**
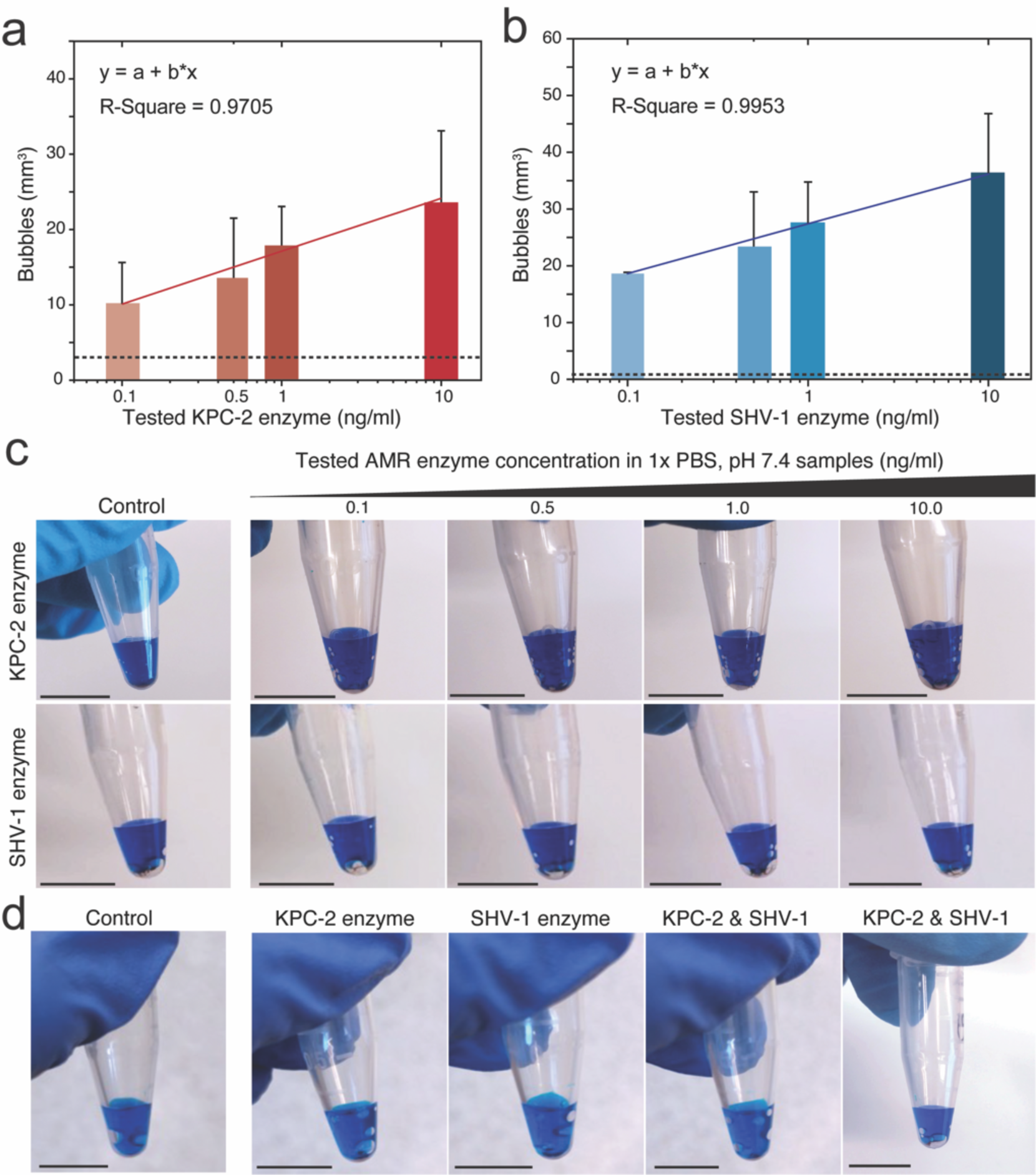
Evaluation of NEIDx system for KPC-2 and SHV-1 detection. (**a**) The change in bubble signal formation in the presence of different concentrations of KPC-2 enzyme (from 0.1 ng/ml to 10 ng/ml). (**b**) The change in bubble signal formation in the presence of different concentrations of SHV-1 enzyme (from 0.1 ng/ml to 10 ng/ml). The dashed lines represent the average value of signal with control samples (no target added). (**c**) Representative images of bubbles formed in the control sample and samples with different concentrations of KPC-2 and SHV-1. (**d**) Representative images of bubbles formed in the control sample, and in the presence of a mixture of KPC-2 and SHV-1 enzymes compared with individual enzyme alone.

We then developed an AI-driven algorithm for imaging and detecting bubbles (i.e., NEIDx Bubble App) to implement the NEIDx approach into a hardware-free cellphone system that allows for rapid qualitative testing of samples. The developed algorithm utilizes a Progressive Web App (PWA) framework (**Figure 5a, b**) that seamlessly integrates trained YOLO (You Only Look Once) and CNN (Convolutional Neural Network) algorithms on a web server[48-50]. The PWA architecture involved the use of the YOLOv8 model to facilitate object detection and Region of Interest (ROI) cropping (**Figure S12**), while the CNN algorithm handled classification based on the presence and absence of bubbles. The YOLOv8n algorithm processes the sample image, and upon identification of the target area of bubble solution, the algorithm automatically crops and forwards the image to CNN that utilizes the MobileNetV2 architecture powered by Google’s TensorFlow deep learning framework (**Figure 6a**). For developing the algorithm, we generated a dataset using KPC-2 spiked into phosphate-buffered saline (PBS) at pH 7.4. The KPC-2 samples varied in enzyme concentration from 0 - 100 ng/ml. The detected lower threshold value was 0.1 ng/ml, which is more sensitive than commercial immunoassay kits for AMR enzyme testing[51]. The resulting dataset comprised 8625 training images and 1931 validation images from 26 videos. Prior to model training, the YOLO component was employed to crop images (**Figure 5a**) followed by training the MobileNetV2 model. The training step yielded 98% training accuracy, 7% training loss, 97% validation accuracy, and 6% validation loss (**Figure S13**). In addition, the NEIDx algorithm was developed with a user-friendly application interface that is easy to follow, consisting of just three simple steps (**Figure 5b**). The app facilitates sample testing by enabling the use of both images that are captured in real-time (**Figure 5b top panel**) and stored images (**Figure 5b bottom panel**) on the system running the application. This user-friendly design enhances accessibility and ensures a seamless experience for users. The performance testing of the developed NEIDx app using spiked-PBS samples demonstrates its proficiency by accurately classifying 93 out of 100 tested sample images (50% negative and 50% positive around the threshold value of 0.1 ng/ml used in the training of the algorithm) (**Figure S14a**), which is significantly improved compared to the model that was generated with MobileNetV2 only (**Figure S14b**). At this threshold, the area under the curve (AUC) was 0.98, with an exact binomial Confidence Interval (CI) ranging from 0.919 to 0.981 as indicated by the Receiver Operating Characteristic (ROC) analysis (**Figure 5c**), confirming the robustness and reliability of the model in classifying samples based on the presence and absence of bubble signal. This was further confirmed with the results of the confusion matrix analysis that showed high accuracy in classifying samples (611 out of 656 of the total number of the tested images) (**Figure S15**). Leveraging the advancements in web technologies, this app runs on a web server combining the advantages of offline functionality with object detection (using YOLO) and image classification capabilities (using CNN known to effectively analyze layers of each tested image with a focus on bubbles as a signal see **Figure S16**), thereby addressing the challenges posed by limited internet connectivity[52, 53]. It is worth noting that the developed NEIDx app is accessible and functional across various devices and operating systems. **Figure S17** shows the developed app running on Android (Motorola XT1575), iOS (Apple iPhone 14 Pro), and Windows (Lenovo IdeaPad S145-15IWL). This broad compatibility enhances the app’s versatility and allows users to seamlessly engage with its features regardless of their preferred device or operating system.

**Figure 5.**
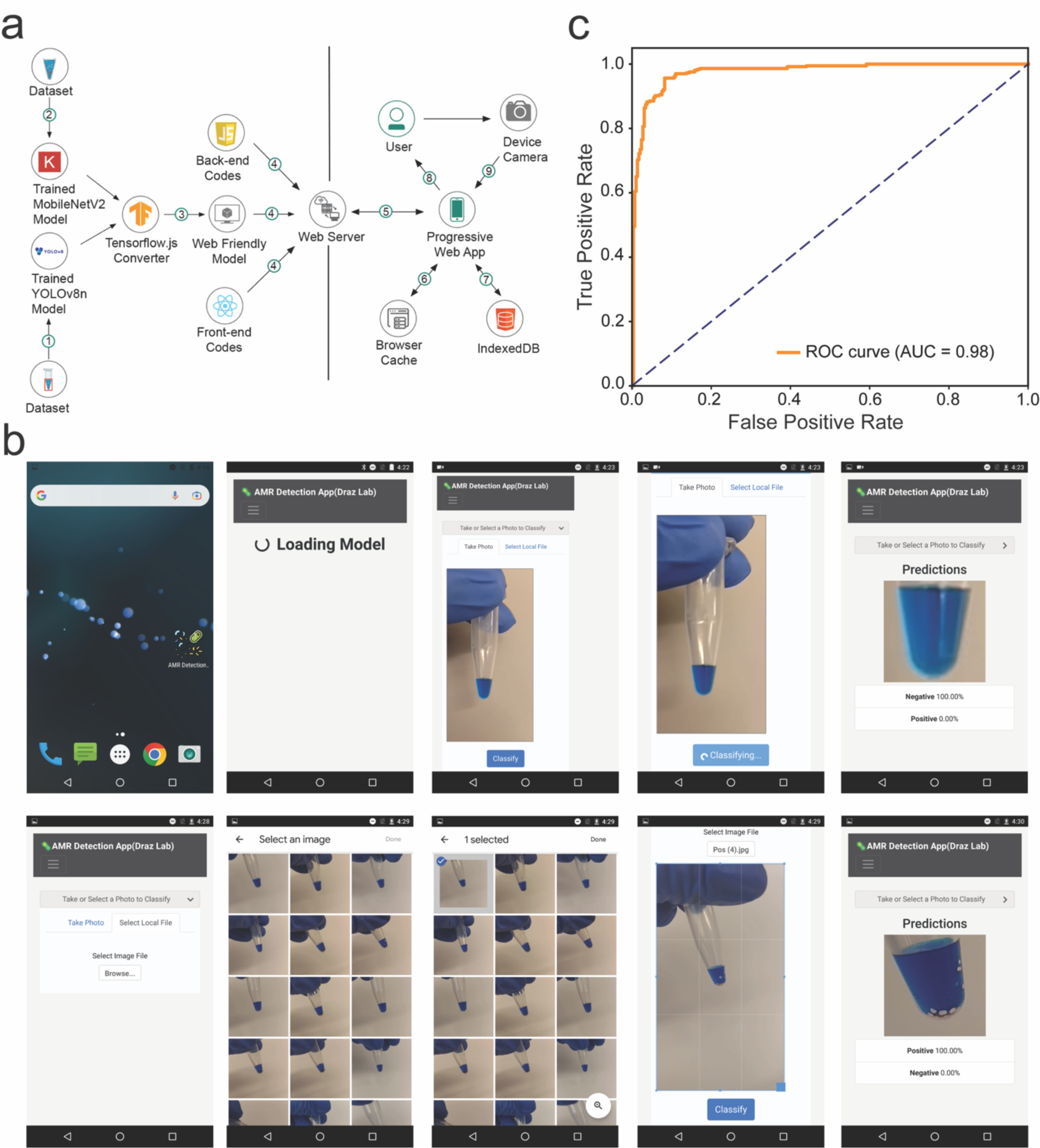
NEIDx Bubble application development and integration for hardware-free sample testing. (**a**) The architectural design of the developed NEIDx system with the Bubble App. (**b**) Bubble detection application interface and data processing. Once a device accesses the AMR progressive web application (PWA) IP address, the necessary files are downloaded to the device and PWA suggests creating a shortcut to the home screen for ease of access. Once the PWA is ready, the user can select between capturing or loading image from device storage. The images loaded into the PWA’s algorithm are processed and classified, and when the process is complete, the result and processed image are displayed on the screen. (**c**) Receiver operating characteristic (ROC) curve analysis of the performance of the NEIDx Bubble application when testing KPC-2 spiked PBS samples. The model was tested with a set of 656 images generated from PBS samples that contain different concentrations of KPC-2 enzymes (0 – 100 ng/ml).

**Figure 6.**
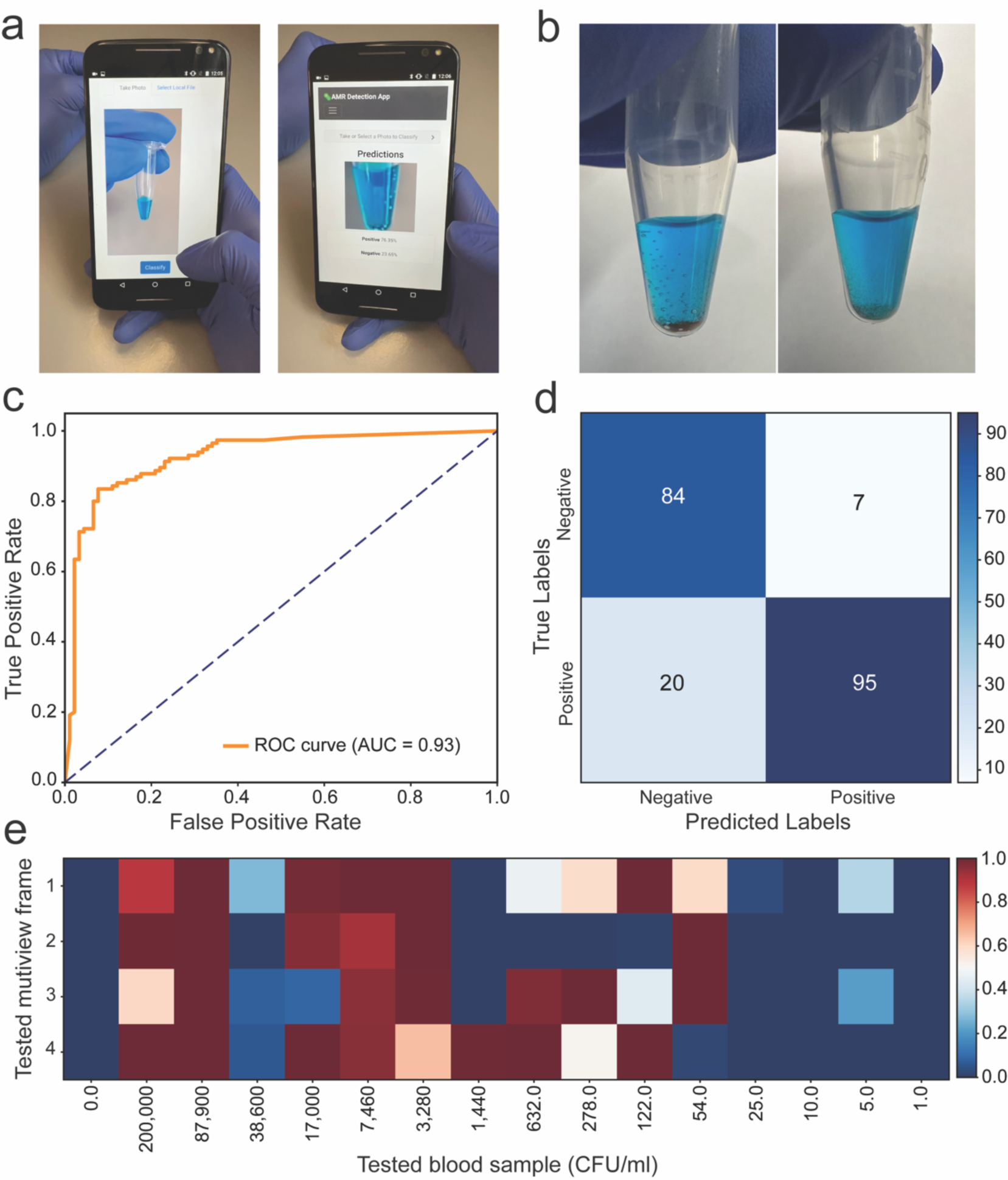
NEIDx system performance in blood sample testing for AMR bacteria. (a) NEIDx Bubble application running on a cellphone system for AMR testing in blood. (b) Representative digital images of samples tested positive (8.79x10^4^ CFU/ml; left) and negative (2 CFU/ml; right) using NEIDx system (i.e., NEIDx integrated with cellphone running the bubble application). (c) Receiver operating characteristic (ROC) curve analysis of the performance of NEIDx system in testing *Klebsiella pneumoniae* spiked blood samples (n = 17). (d) The confusion matrix analysis of bacteria spiked blood samples (n = 17). (e) Heatmap of the probability value of sample measured by the NEIDx system for different bacteria concentrations spiked in blood (n = 17). The AMR sample with ≥ 0.1 ng/ml is considered positive (red in color; probability value ≥0.5) and sample with <0.1 ng/ml is considered negative (blue in color; probability value < 0.5).

### Blood sample testing using NEIDx system

To further confirm the potential of the developed NEIDx system for AMR testing in biological samples, we prepared a set of whole blood spiked samples with a range of *Klebsiella pneumoniae* (1 – 1x10^5^ CFU/ml) (**Figure 6a-e**). Each of the samples was lysed and incubated with MBs for 20 minutes then allowed to react with Pt-nanoprobes for 15 minutes followed by a washing step and the addition of peroxide solution. **Figure 6a** captures the NEIDx system in action as it evaluates the bubble results of the tested samples. Images acquired by the developed AI algorithm during the testing process, for both positive and negative samples, are visually showcased in **Figure 6b**. The results produced by the NEIDx system indicated a direct correlation between the tested concentration of lysed bacteria and the developed bubble signal (**Figures S18, 19**). The ROC analysis showed that target KPC-2 enzyme concentration of 0.1 ng/ml yields an optimum sensitivity of 82.61% with a CI of 79.7 – 85.5% and a specificity of 92.31% with a CI of 90.3 – 94.3%. At this threshold, the area under the curve (AUC) was 0.93, with an exact binomial CI ranging from 0.91 – 0.95% (**Figure 6c**). The confusion matrix analysis revealed that NEIDx correctly classified 86.89% of all trials conducted for blood sample testing, demonstrating exceptional performance with a precision of 0.93 (**Figure 6d**). The F1 Score of 0.88 indicates a well-balanced trade-off between precision and recall. Notably, the model exhibits a specificity of 0.92, crucial for minimizing false positives. The Matthews correlation coefficient (MCC) reaches 0.74, signifying strong agreement between predicted and actual classifications of the tested samples using NEIDx. These metrics underscore the system’s robust capability for precise classification, unaffected by variations in imaging angles or the orientation of the tested sample, which may vary from one user to another (**Figure S20**). Furthermore, the qualitative results as probability values, for being positive (≥0.1 ng/ml) or negative (<0.1 ng/ml), obtained by the NEIDx system compared with the tested concentrations of *Klebsiella pneumoniae* in spiked blood samples are represented in a heatmap plot shown in **Figure 6e**. These findings offer intricate insights into the robust performance and diagnostic accuracy of the NEIDx system for AMR testing, particularly in whole blood samples.

## CONCLUSIONS

AMR presents a growing threat to global public health, demanding innovative diagnostic tools for rapid identification and characterization of resistant pathogens. Our AI-driven system, NEIDx, addresses this need by streamlining AMR testing directly from blood samples. Leveraging AI and nanotechnology, NEIDx eliminates bulky hardware and extensive automation, enabling the rapid detection of AMR-associated enzymes through catalytic nanoparticle-based technology. The results reveal NEIDx’s exceptional sensitivity (0.98) and specificity (0.93) in detecting clinically relevant AMR enzymes, such as KPC-2 and SHV-1, in blood samples. Furthermore, NEIDx’s versatility allows simultaneous detection of multiple target enzymes, enhancing its applicability across diverse healthcare settings. The development of an AI-driven algorithm for image analysis, integrated into a user-friendly application, ensures seamless and reliable sample testing, even in resource-limited environments. With a training accuracy of 97% and classification accuracy of 93%, the algorithm reliably detects the presence of AMR-associated enzymes. Validation studies using whole blood samples further underscore NEIDx’s potential for real-world application, demonstrating impressive sensitivity, specificity, and precision. NEIDx represents a significant advancement in POC diagnostics for AMR, offering a transformative solution to combat the urgent global challenge. Its user-friendly interface, hardware-free operation, and exceptional performance position NEIDx as a promising tool for revolutionizing AMR testing and improving public health outcomes worldwide.

## MATERIAL AND METHODS

### Preparation and characterization of NEIDx magnetic beads

Antibody oxidation and coupling protocols were previously described.^14^ Briefly, anti-KPC-2 and anti-SHV-1antibodies were prepared in 0.1 M sodium acetate (pH 5.5) and 10 mM sodium metaperiodate (pH 5.5) mixture and incubated on ice for 20 min in the dark. The oxidized antibodies then washed with 0.01 M PB before activated with PDPH crosslinker for 1 hour. PDPH activated antibodies were incubated with thiolated MBs for 4 hours at 4 °C on an orbital shaker. Following the incubation of MB with antibodies, the excess solution (supernatant) was removed, and MB were washed in PB on a magnetic separator. The prepared beads and the efficiency of the coupling reaction were characterized using UV-vis, fluorescence spectroscopy, and SDS-PAGE analysis techniques.

### Preparation and characterization of NEIDx catalytic nanoprobes

Catalytic nanoprobes that specifically recognize KPC-2 and SHV-1 enzymes captured on NEIDx beads were prepared using platinum nanoparticles (PtNPs). The preparation protocol starts with PtNPs synthesis followed by antibody coupling to the surface of PtNPs using PDPH crosslinker. PtNPs were synthesized using a modified method previously reported. Aqua regia and ultrapure water was used to clean all glassware. A 0.2% solution of chloroplatinic acid hexahydrate was added to boiling DI water followed by the addition of a solution containing 1% sodium citrate and 0.05% citric acid, and a quick injection of a freshly prepared 0.08% sodium borohydride solution containing 1% sodium citrate and 0.05% citric acid. After 10 min of reaction, the NP solution was cooled down to RT. The freshly prepared PtNPs were first activated using PDPH crosslinker, then allowed to pair with the oxidized antibody for 1hour at room temperature. The prepared Pt-nanoprobes were washed and characterized using UV-vis spectroscopy, FT-IR spectroscopy, transmission electron microscopy, and agarose gel electrophoresis analysis techniques.

### NEIDx testing and performance evaluation

To evaluate the sensitivity of the NEIDx system, dilutions of KPC-2 and SHV-1 enzyme stocks ranging from 0.0 ng/ml to 10 ng/ml were prepared in 1x phosphate buffer (PB) pH 7.4. Aliquots (10 μL) of the tested concentrations were tested using the NEIDx system. The targeted enzymes are first incubated with NEIDx beads for 10 – 15 minutes before being washed and mixed with NEIDx Pt-nanoprobes for an additional 10 minutes at room temperature. The formed catalytic immunocomplex was then mixed with 100 μL of the peroxide-containing colored solution (100 μL of glycerol, 670 μL of DI water, 200 μL of 30% (v/v) H_2_O_2_, and 30 μL of blue colored food dye) and tested for the presence of oxygen bubbles. A similar protocol was followed for the multiplex testing analysis using a mixture of NEIDx beads and nanoprobes that specifically recognize the targeted mix of AMR enzymes of KPC-2 and SHV-1. The generated bubble area was measured manually using ImageJ tool version 1.53k.

### NEIDx AI-enabled application

A dataset of 10556 prelabeled images, organized in two groups around a threshold concentration of 0.1 ng/ml as positive (≥ 0.1 ng/ml) and negative (< 0.1 ng/ml), was utilized for NEIDx app training and validation. For object detection, a subset of 1400 images from the training dataset were employed for YOLOv8n model training, and a subset of 100 images was used for testing. The fully developed YOLOv8n model was utilized to generate a dataset of cropped images for the training and testing of the CNN model of MobileNetV2. The algorithm was trained with this data set with batch size 32, number of epochs 100, and learning rate 0.001.

To develop a PWA to support the NEIDx system, we utilized a standard development environment, comprising a high-performance workstation equipped with modern web development tools, including HTML, CSS, and JavaScript frameworks. The application’s frontend was primarily developed using the React library, ensuring a highly responsive and user-friendly interface. The interface was crafted using HTML and CSS, with interactive elements implemented using JavaScript and React components. Responsive design principles ensure compatibility across various types of devices and operating systems. The developed PWA uses the tested and trained YOLOv8n and MobileNetV2 algorithms to detect and classify the assay results. Leveraging the versatility of web technologies, this PWA combines the advantages of offline functionality with object detection and image classification capabilities, thereby addressing the challenges posed by limited internet connectivity. To enable offline functionality, the models were saved in locally stored IndexedDB databases after first access to the website, and in offline situations, models are pulled from this database. The frontend and backend data were stored on cache storage to enable the user interface and backend codes to work seamlessly in offline situations by NEIDx users.

### NEIDx performance in testing blood samples

We evaluated the performance of the developed NEIDx system in testing *Klebsiella pneumoniae*-spiked blood samples (n = 17) to simulate real-world samples. Fresh blood from healthy donors, commercially obtained from Research Blood Components, LLC., was spiked with known concentrations of a resistant *Klebsiella pneumoniae* strain (0 – 10^5^ CFU/ml) and blindly tested using NEIDx system. Aliquots of 200 µL were taken from the spiked whole blood samples, lysed with sonication (5 minutes) then mixed with NEIDx beads and nanoprobes and the generated bubbles signal was evaluated using the NEIDx app running on an optical system with a camera, such as cellphone or tablet devices.

## Data Availability

All data produced in the present work are contained in the manuscript

